# Evaluating COVID-19 Public Health Messaging in Italy: Self-Reported Compliance and Growing Mental Health Concerns

**DOI:** 10.1101/2020.03.27.20042820

**Authors:** Soubhik Barari, Stefano Caria, Antonio Davola, Paolo Falco, Thiemo Fetzer, Stefano Fiorin, Lukas Hensel, Andriy Ivchenko, Jon Jachimowicz, Gary King, Gordon Kraft-Todd, Alice Ledda, Mary MacLennan, Lucian Mutoi, Claudio Pagani, Elena Reutskaja, Christopher Roth, Federico Raimondi Slepoi

## Abstract

**Purpose:** The COVID-19 death-rate in Italy continues to climb, surpassing that in every other country. We implement one of the first nationally representative surveys about this unprecedented public health crisis and use it to evaluate the Italian government’ public health efforts and citizen responses.

**Findings:** (1) Public health messaging is being heard. Except for slightly lower compliance among young adults, *all* subgroups we studied understand how to keep themselves and others safe from the SARS-Cov-2 virus. Remarkably, even those who do *not trust the government*, or *think the government has been untruthful* about the crisis believe the messaging and claim to be acting in accordance. (2) The quarantine is beginning to have serious negative effects on the population’s mental health.

**Policy Recommendations:** Communications should move from explaining to citizens that they should stay at home to what they can do there. We need interventions that make staying following public health protocols more desirable, such as virtual social interactions, online social reading activities, classes, exercise routines, etc. — all designed to reduce the boredom of long term social isolation and to increase the attractiveness of following public health recommendations. Interventions like these will grow in importance as the crisis wears on around the world, and staying inside wears on people.

## 1. Introduction

At the request of government officials from the country hardest hit by the COVID-19 crisis, we evaluate public understanding of Italian government public health messaging about the crisis. We do this via a nationally representative survey of 3,452 Italian adults between March 18th 2020 and March 20th 2020. A statement of ethical approval appears in Appendix A.

We draw five main conclusions and present evidence for each in the following five sections. First, most demographic groups, especially the elderly, believe in and follow health measures (Section 2). Second, even skeptics about the seriousness of the disease and of the government’s messaging, truthfulness, and abilities, still largely believe the messaging and report being in compliance with it (Section 3). Third, most people, especially the elderly and infirm, leave home only for reasons deemed “essential” (Section 4). Fourth, the population is experiencing high levels of anxiety, especially vulnerable groups (Section 5). And finally, nudges to improve attitudes and compliance that are already near their maxima have little effect (Section 6). Methodological details, including survey questions, appear in Appendix B.

## 2. Most demographic groups, especially the elderly, believe in and follow health measures

We now present two pairs of figures describing *beliefs about public health measures* (Figures 1 and 2) and *compliance with recommended health practices* (Figures 3 and 4). The first figure in each pair gives overall results, and the second is broken down by demographic group.

**Figure 1:**
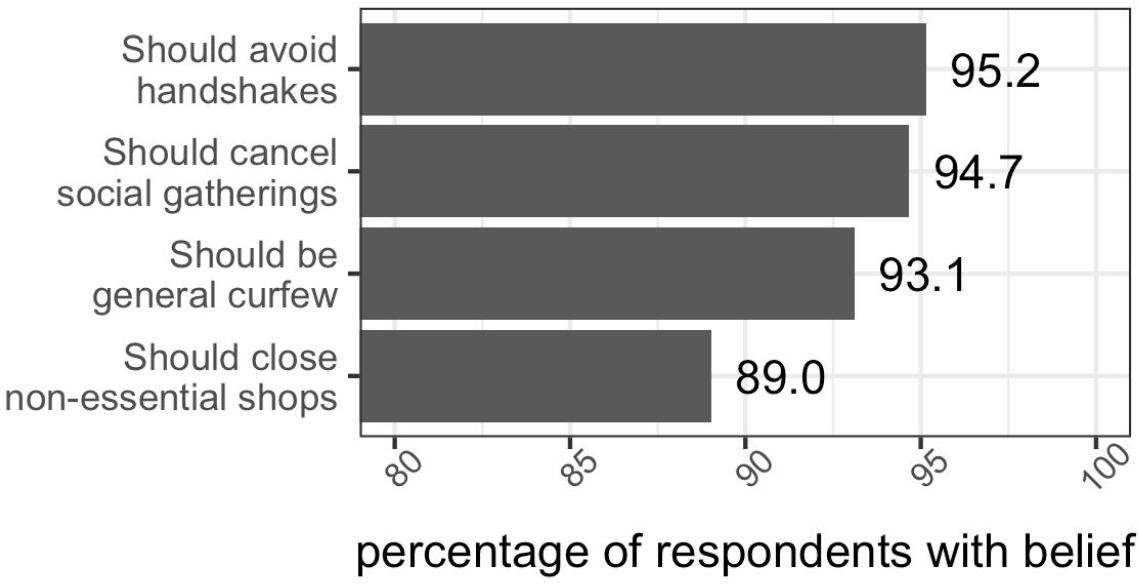
Respondents’ Beliefs about Public Health Measures. Bars and numbers correspond to percentages of “yes” responses to questions in item 3.1 in the Appendix B.

**Figure 2:**
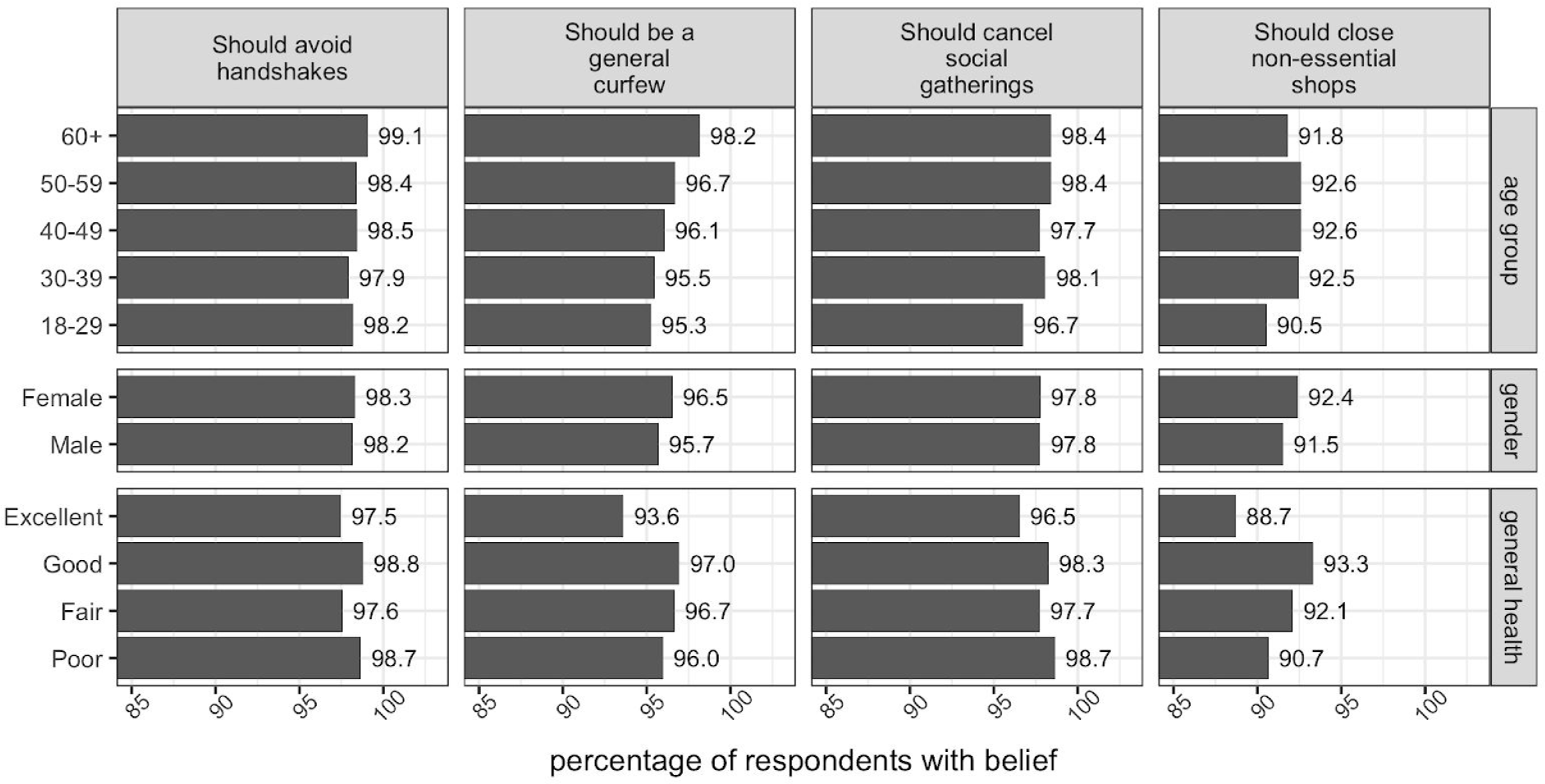
Respondents’ Beliefs about Public Health Measures: By Demographic Group. Bars and numbers correspond to percentages of “yes” responses to questions in item 3.1 in Appendix B.

**Figure 3:**
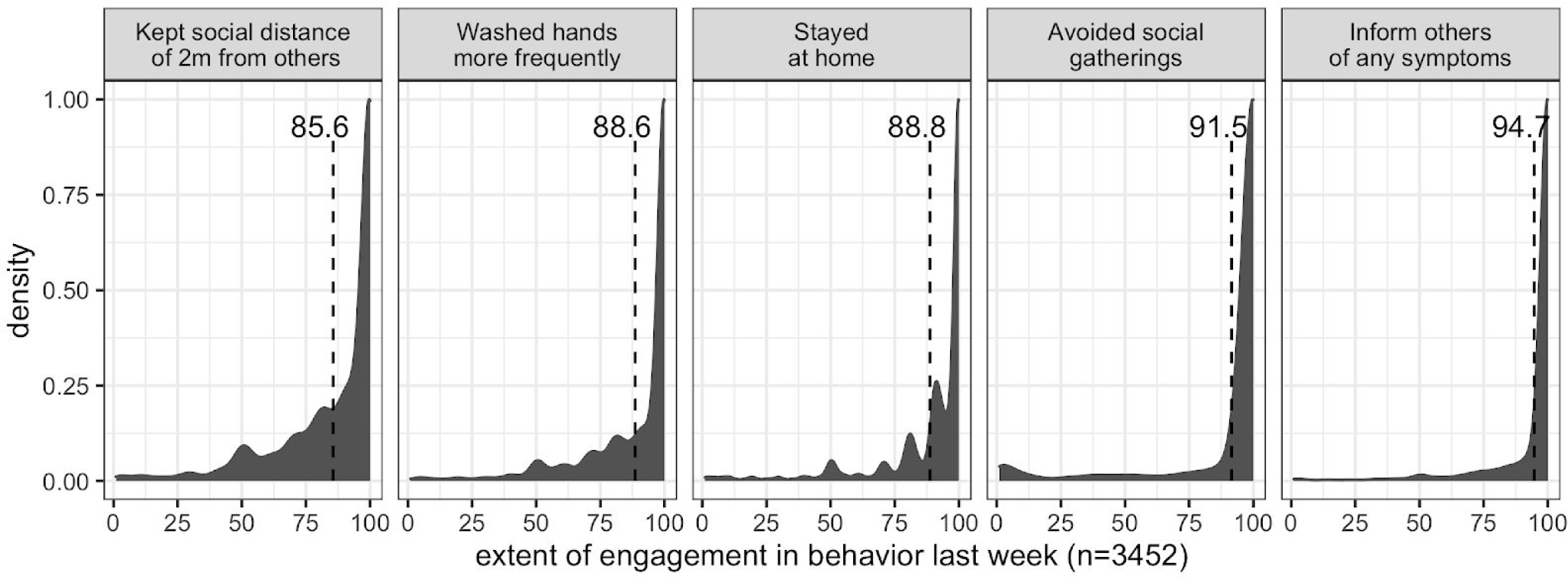
Respondents’ Compliance with Health Practices in the Previous Week. Histograms correspond to responses to item 1 (“*In the past week, on a scale from 1-100, to what extent did the following describe your behavior?”*) in the Appendix B. Dashed lines show average score.

**Figure 4:**
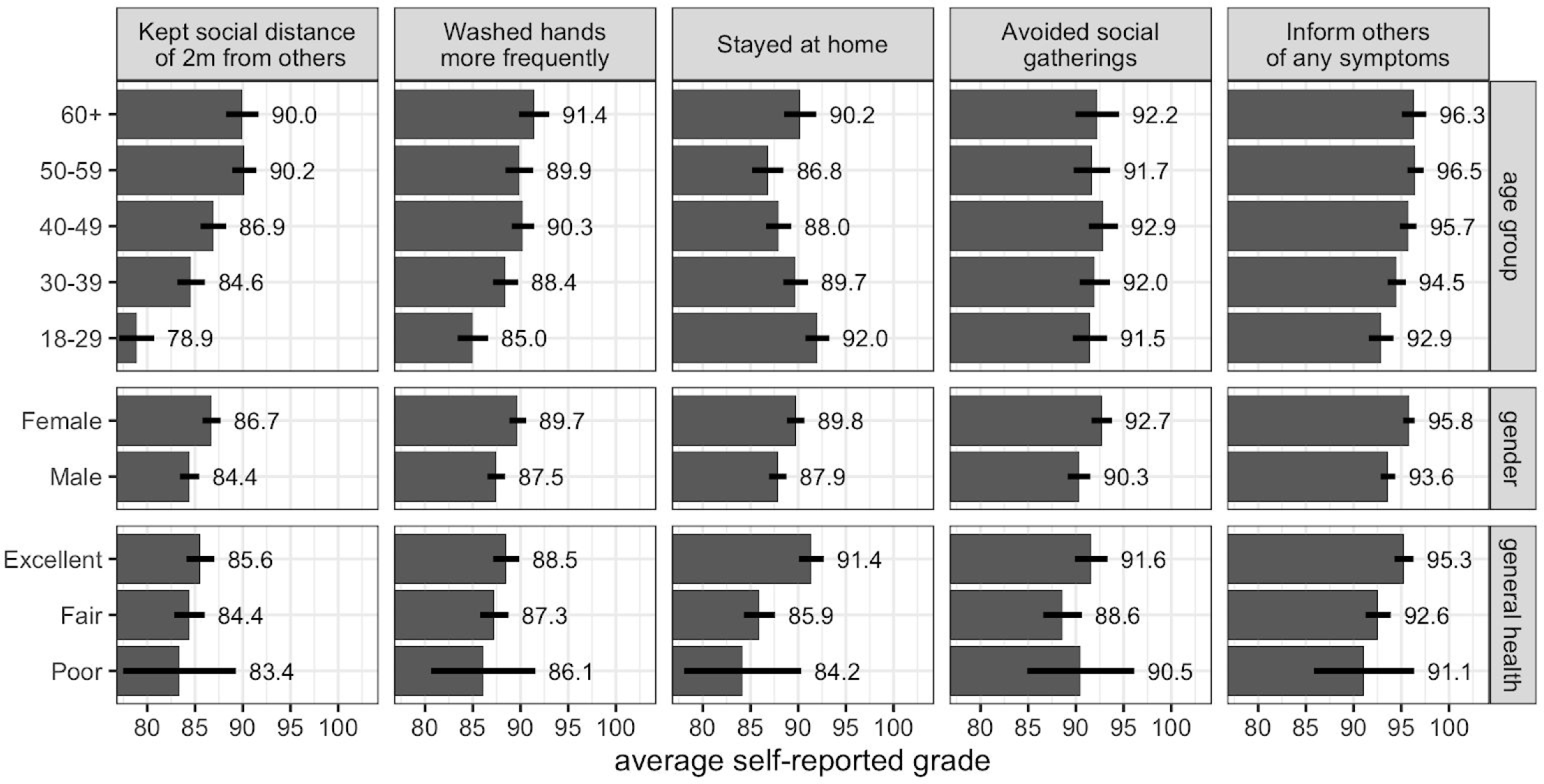
Respondents’ Compliance with Health Practices in the Previous Week: By Demographic Group. Bars and numbers indicate average self-reported response (out of 100) for respondents belonging to each group on the left hand side (with 95% Confidence Intervals). Plots correspond to responses to item 1 (“*In the past week, on a scale from 1-100, to what extent did the following describe your behavior?”*) in Appendix B.

**Figure 4:**
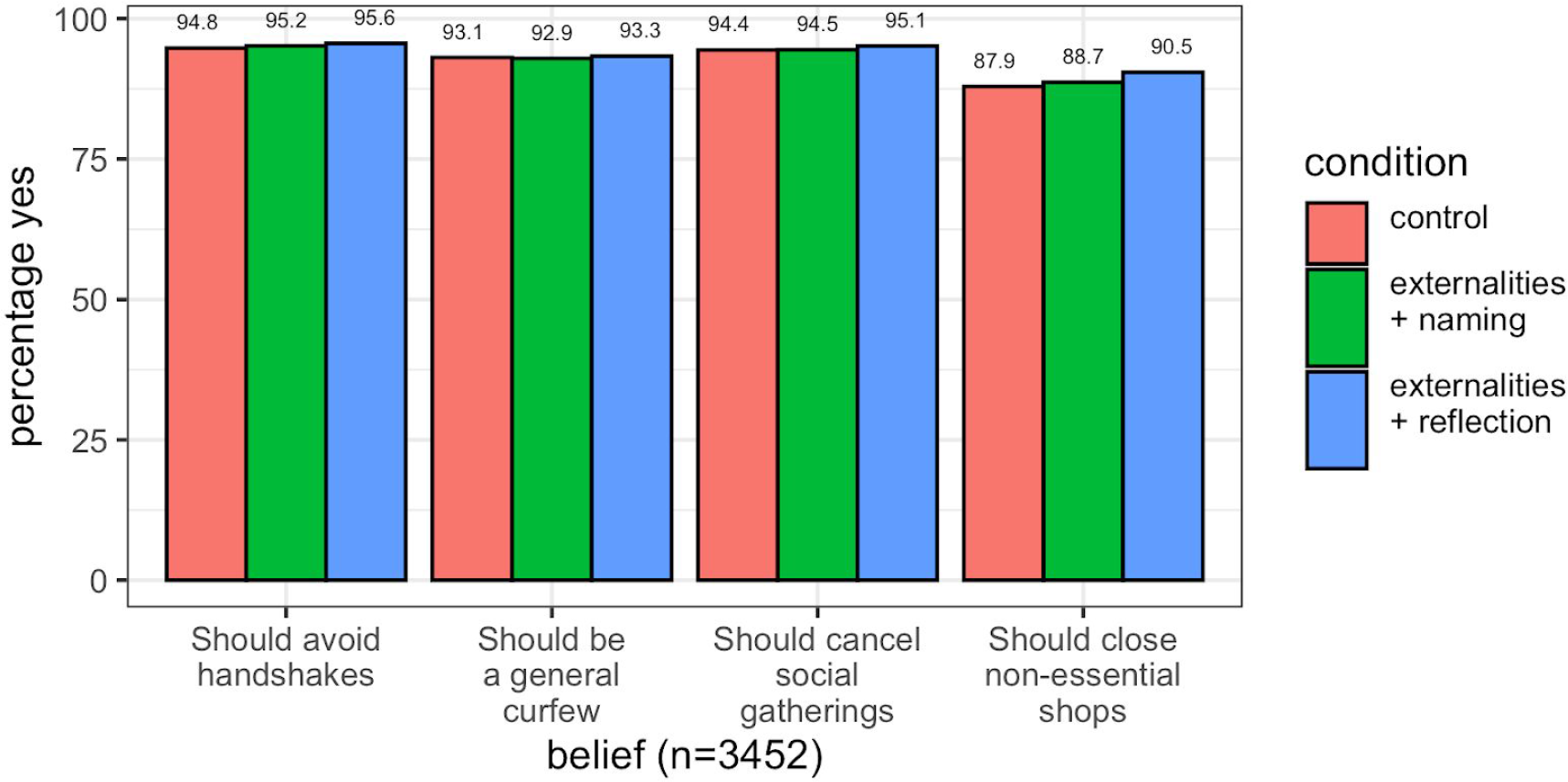
Information Treatments Did Not Increase Pro-Health Beliefs. Each bar indicates the percent of respondents in each treatment condition who answered yes to each question.

What does the Italian public believe about recommended health measures regarding the COVID-19 crisis? Figure 1 shows that our subjects are appropriately cautious: nearly 100% endorse four recommended public health measures (handshake avoidance, social gathering avoidance, non-essential activities curfew, and non-essential shop closure). Of the four, the curfew and shop closure are slightly less popular, but an 89% endorsement rate is still remarkable. Compared with decades of advice to reduce smoking, drinking, and lack of exercise, these levels are very high.

All demographic groups strongly agree with these four measures, though there is consistent (small) disagreement amongst respondents reporting that they are in “excellent” health (Figure 2). When asked to reflect in writing about the social costs of reneging on the quarantine, nearly *all* respondents express agreement and emphasize collective responsibility. A representative (open text) reflection from the entire respondent sample reads^3^:

> *“I believe that EVERYONE should stay at home, leaving only for the bare minimum and taking security measures. Only in this way can we solve and solve this enormous problem in the shortest possible time*.*”*

We note that older respondents (1) consistently express worry or anxiety and (2) perceive that <u>others</u> in the public are reneging on their duty. A representative reflection from the 60+ subgroup reads^4^:

> *“I fully understand everything that is happening, so in spite of myself, I stay at home, but I would like this to include all those people who unfortunately continue to go out, especially the young, even every day! I think the longer we stay home, the sooner we get out!”*

We turn from respondents’ beliefs to their actual behavior. Figure 3 shows that a majority of respondents report that they have been strongly complying with recommended health practices.

Figure 4 shows that seniors (60+), women, and healthier individuals say they are more compliant with most health practices. Relative to others, the young defect from social distancing and hand-washing in a statistically significant sense. Poor-health individuals are notably less compliant with staying at home, but with high variance: the reason may be their frequency of hospital and pharmacy visitation (see Figure 8).

Given the public salience of these health measures, it is likely that self-reports suffer from social desirability bias. Future studies should validate the compliance rates shown here with real-world observational data.

## 3. Skeptics still mostly believe the messaging and say they’re behaving accordingly

Public opinion during this crisis is not entirely positive. Though 78% of respondents believe social distancing is an effective health measure, only 62% think the government’s reaction to the crisis is appropriate (rather than extreme or insufficient) and 36% think the (rest of the) public’s reaction is appropriate. Additionally, a slim majority of 58% believe the government has been truthful regarding information about the crisis, and only 51% of the public say they trust the Italian government to take care of them in a crisis. Are these skeptics – about health measure efficacy, the public, or the government – defecting from public health protocols?

The majority of skeptics^5^ endorse public health protocols, though with exception (red bars in Figure 5). More than 80% within each skeptic group (i.e., low trust of government, believe information is untruthful, believe social distancing is ineffective) hold pro-health beliefs – except for those who believe the government or public is over-reacting to the virus.

**Figure 5:** Skeptics’ (Red) Beliefs about Public Health Measures. Bars and numbers correspond to percentages of “yes” responses to questions in item 3.1 in Appendix B for respondents belonging to each opinion group on the left hand side. Opinion groups membership constructed by trichotomizing responses to the 5-point questions in item 3.4. Highlighted red bars correspond to skeptic groups.

**Figure 5:**
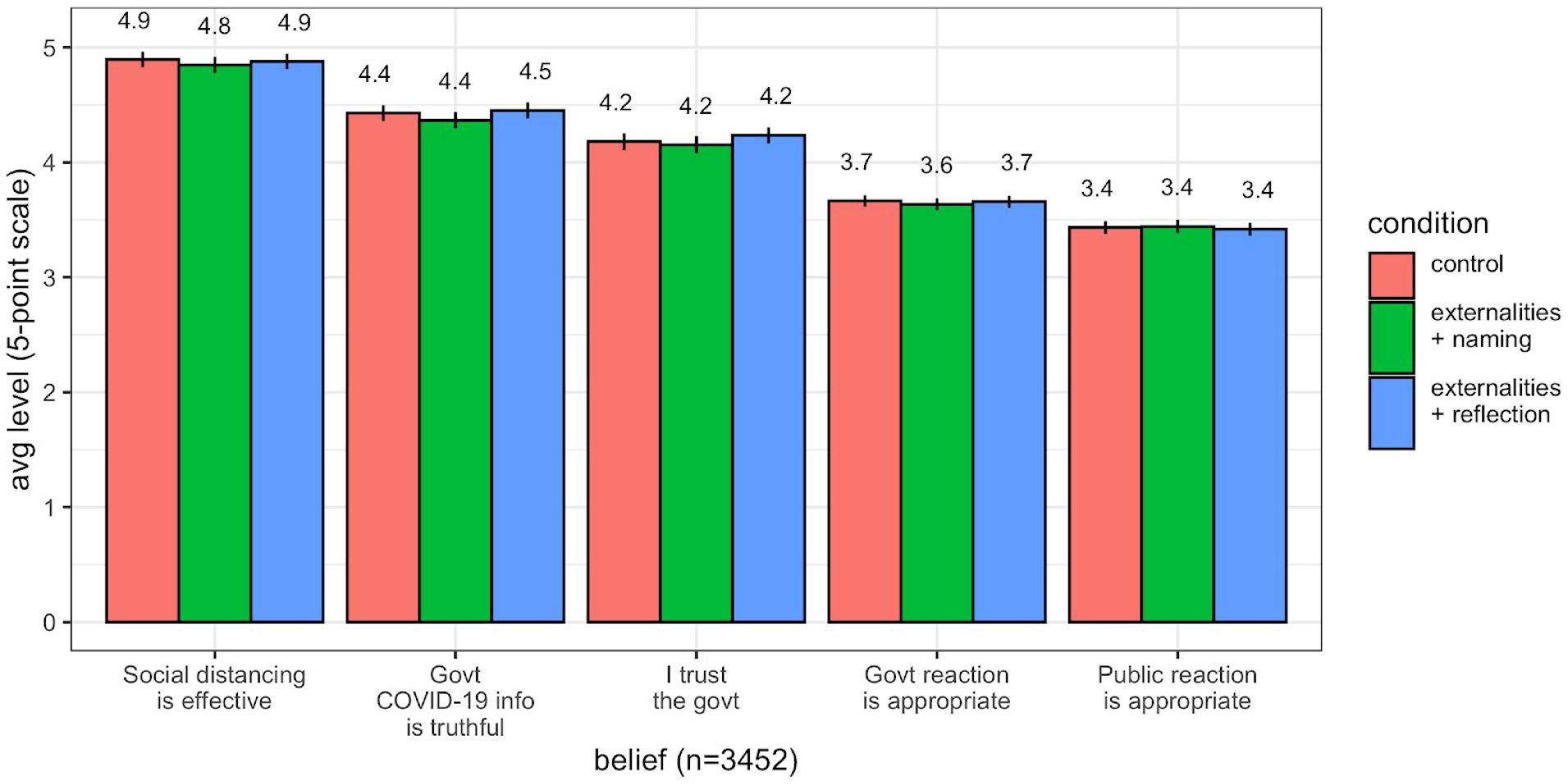
Information Treatments Did Not Decrease Skepticism. Each bar plot indicates the average of responses (out of 5 points) given to each question (95% confidence interval).

Skeptics about the severity of the crisis who were asked to reflect in writing about the social costs of reneging on the quarantine sometimes say that the government is “*exaggerating*” or that “*the economy and the nation are affected*”. However, many still express a willingness to cooperate. As one respondent who thought government reaction was extreme responded^6^:

> *“It’s all just media. Even last year 12 thousand flu deaths and nobody spoke about it. Anyway I carry out the orders even if I don’t understand them. Surely Conte is not commanding but someone else”*

These same trends hold in behavior: remarkably, those who distrust the government, or believe the government may not be disseminating fully correct information, on average, report compliance scores between 82% and 92%. Unsurprisingly, skepticism in social distancing reflects in lower social distancing compliance, though average score remains nearly 80%. Those who believe reactions are over-blown were least compliant in the past week on all behaviors, but even these are with scores consistently above 70%. We do not observe direct behaviors other than through self-reports, and so following this group may be worthwhile if better measures become available.

## 4. Most people, especially the elderly and sick, leave home only for “essential” reasons

We now examine compliance with the quarantine. In particular, we examine *reasons for going out* (Figures 5 and 6), and *negative aspects of staying home* (Figures 7 and 8). The first figure in each pair gives overall results, and the second is by demographic group.

**Figure 6:** Skeptics’ (Red) Respondents’ Compliance with Health Practices in Previous Week. Bars and numbers indicate average self-reported response (out of 100) for respondents belonging to each opinion group on the left hand side (with 95% Confidence Interval). Plots correspond to responses to item 1 (“*In past week, on a scale from 1-100, to what extent did the following describe your behavior?”*) in Appendix B. Opinion groups membership constructed by trichotomizing responses to the 5-point questions in item 3.4. Highlighted red bars correspond to skeptic groups.

**Figure 7:**
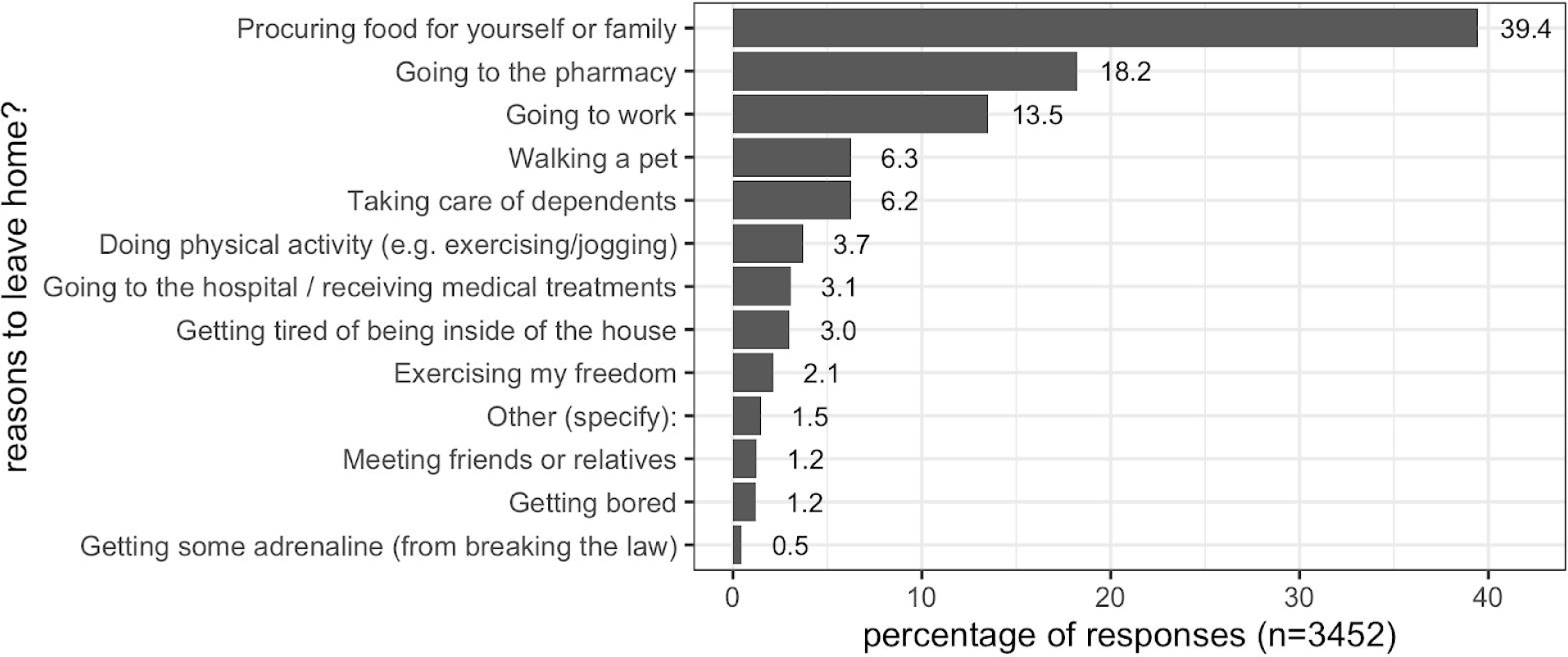
Non-Compliers’ (62% of Sample) Reasons Cited for Leaving Home. Plots correspond to checkboxes picked for question in item 4 in Appendix B: “*What are the reasons personally for you to leave your home (check all that apply)?”*. On average, respondents picked 2 reasons.

**Figure 8:**
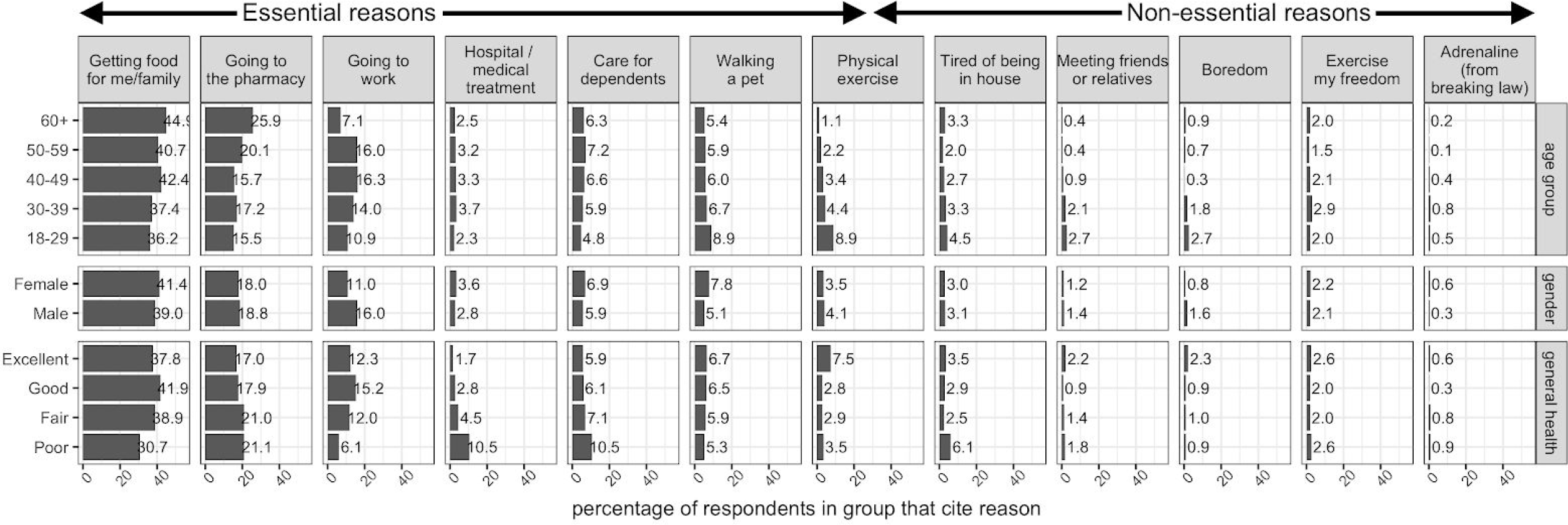
Non-Compliers’ (62% of Sample) Reasons Cited for Leaving Home: By Group. Plots correspond to responses picked for question in item 4 in the survey design appendix: “*What are the reasons personally for you to leave your home (check all that apply)?”*. On average, respondents picked 2 reasons.

Sixty-two percent of all respondents say they <u>need</u> to leave home in the next 5 days. Why are these respondents defecting from the quarantine? Figure 7 examines respondents’ reasons for not staying home in the next 5 days. Respondents most often cite leaving home for *essential* reasons (e.g., procuring food, retrieving medicine) rather than *non-essential* reasons (e.g., getting bored, meeting friends). Of those who express a need to leave, 95% cite at least one essential reason while only 18% cite any non-essential reasons.

Figure 8 shows that, generally, those needing to leave for *essential* reasons are more often older while those leaving for increasingly *non-essential* reasons are (slightly more often) younger people below the age of 30. Older people say they are defecting to procure groceries; the middle-aged to go to work; the less healthy and elderly to receive medical treatment.

Respondents broadly cite the loss of mental health needs – freedom, stimulation, fresh air, and physical exercise – as the most common negative experiences associated with the quarantine (Figure 9).

**Figure 9:**
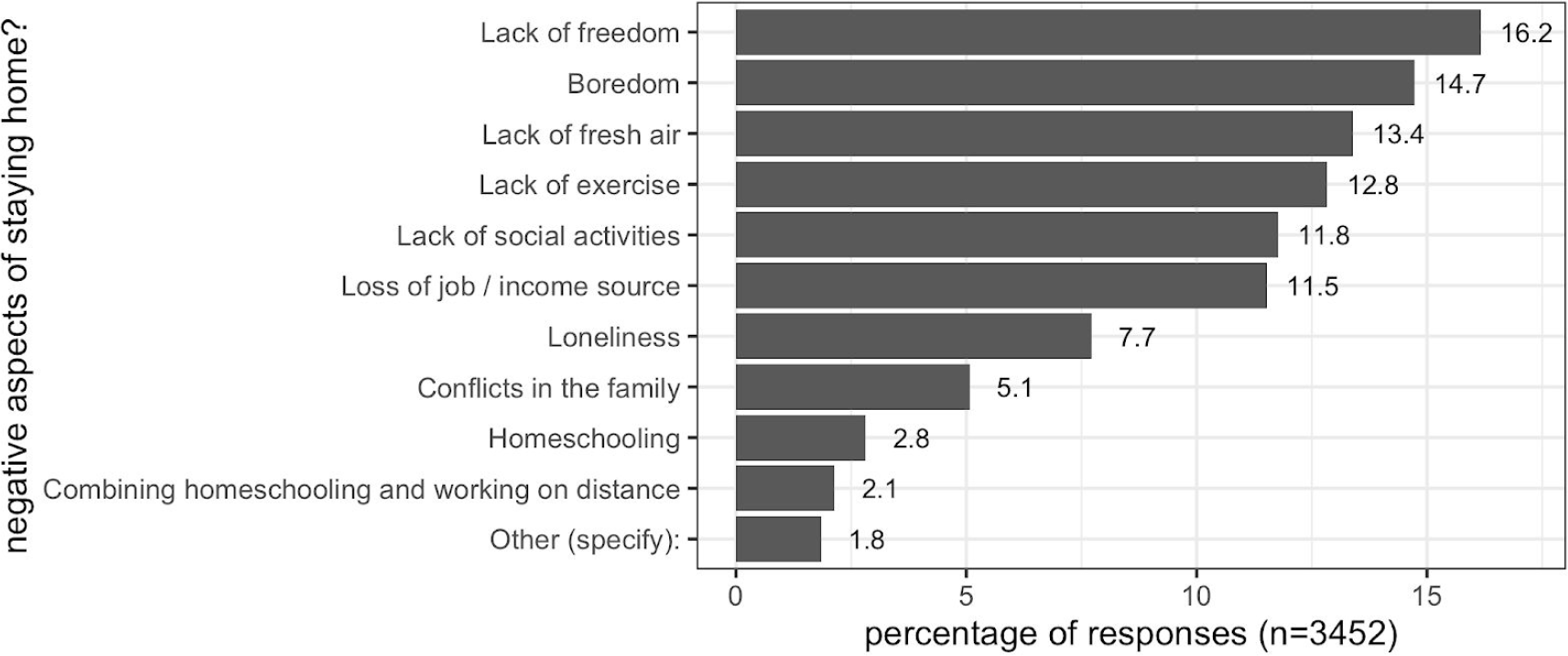
Negative Aspects Cited by all Respondents of Staying Home. Plots correspond to responses to item 4 in Appendix B: “*What are the main negative sides of complying with stay home requirement (check all that apply)?”*. On average, respondents picked 2 reasons.

Still, there are group-specific negative experiences (Figure 10). Younger people more often say they are struggling with increased boredom and family conflict. Meanwhile, vulnerable groups like the elderly and the health-compromised more consistently cite loneliness relative to others.

**Figure 10:**
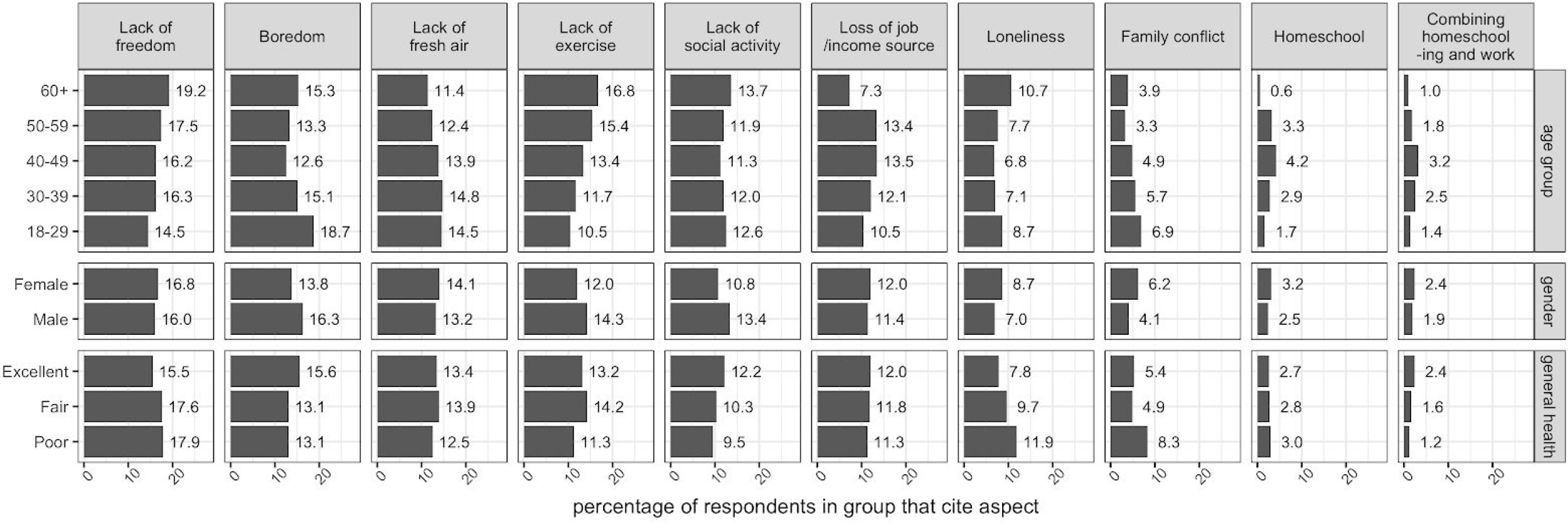
Negative Aspects Cited by all Respondents of Staying Home: By Group. Plots correspond to responses to item 4 in the survey design appendix: “*What are the main negative sides of complying with stay home requirement (check all that apply)?”*. On average, respondents picked 2 reasons.

Those who are likely working parents (aged 40-49) more consistently cite economic distress and struggles with home-schooling and work than other groups (which, of course, might well have been the case if we had a control group from before the crisis). Overall, this confirms that different demographic groups are struggling with different aspects of quarantining. Going forward, these groups might benefit from personalized “morale-boosting” interventions.

## 5. High levels of anxiety in the population, particularly amongst vulnerable groups

The average level of anxiety surrounding the crisis in the population is high: none of our respondents reported being completely without anxiety (Figure 11).

**Figure 11:**
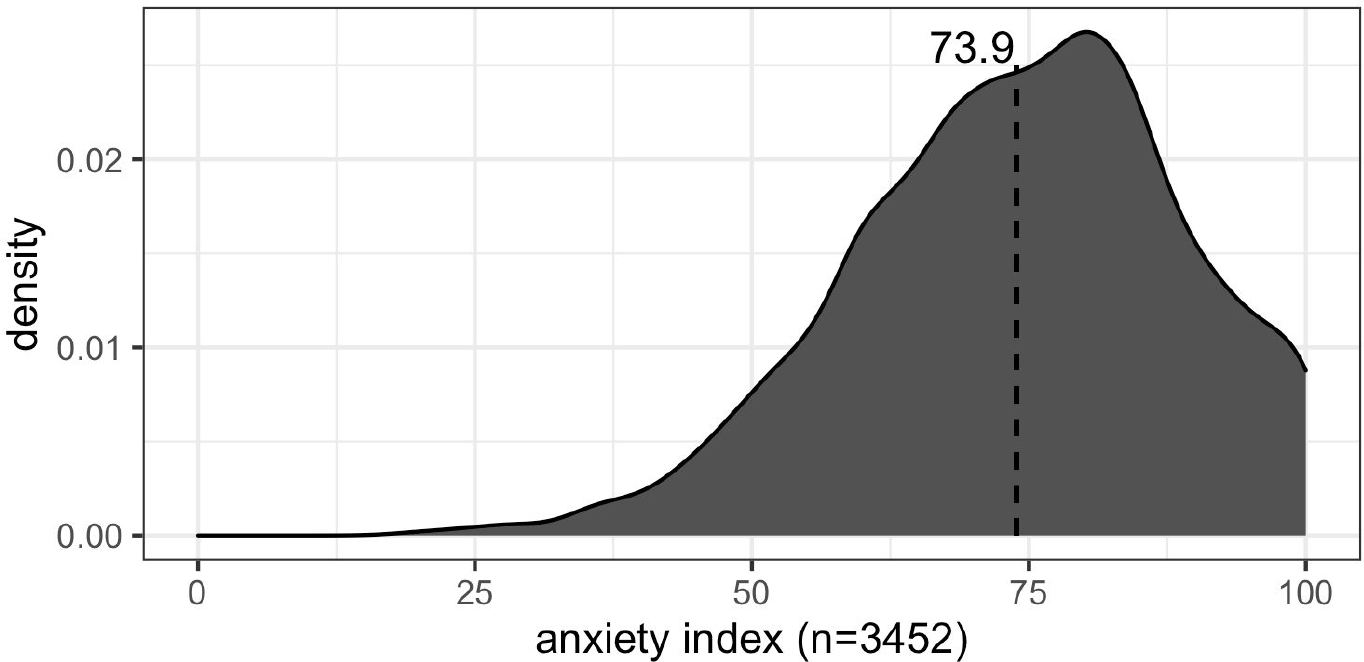
Respondents’ Overall Coronavirus-related Anxiety Level. Histogram indicates distribution of respondents’ average of 5 responses to 5-point questions: (1) “*I am nervous when I think about current circumstances* (2) “*I am calm and relaxed”* (3) “*I am worried about my health”* (4) “*I am worried about the health of my family members”* (5) *“I am stressed about leaving my house”*.

Who is most anxious about the circumstances right now? Figure 12 shows that the most anxious relative to their group are: women, respondents with poor health, and older adults. Although anxiety might be driving healthy behaviors and pro-health beliefs described above, additional anxiety due to ones’ vulnerable status is likely to negatively impact long-term mental health. It is possible that increased messaging may not inform but rather exacerbate anxiety for inherently vulnerable groups like seniors and the immunocompromised. Despite these differences, the overall levels are very high for every group.

**Figure 12:**
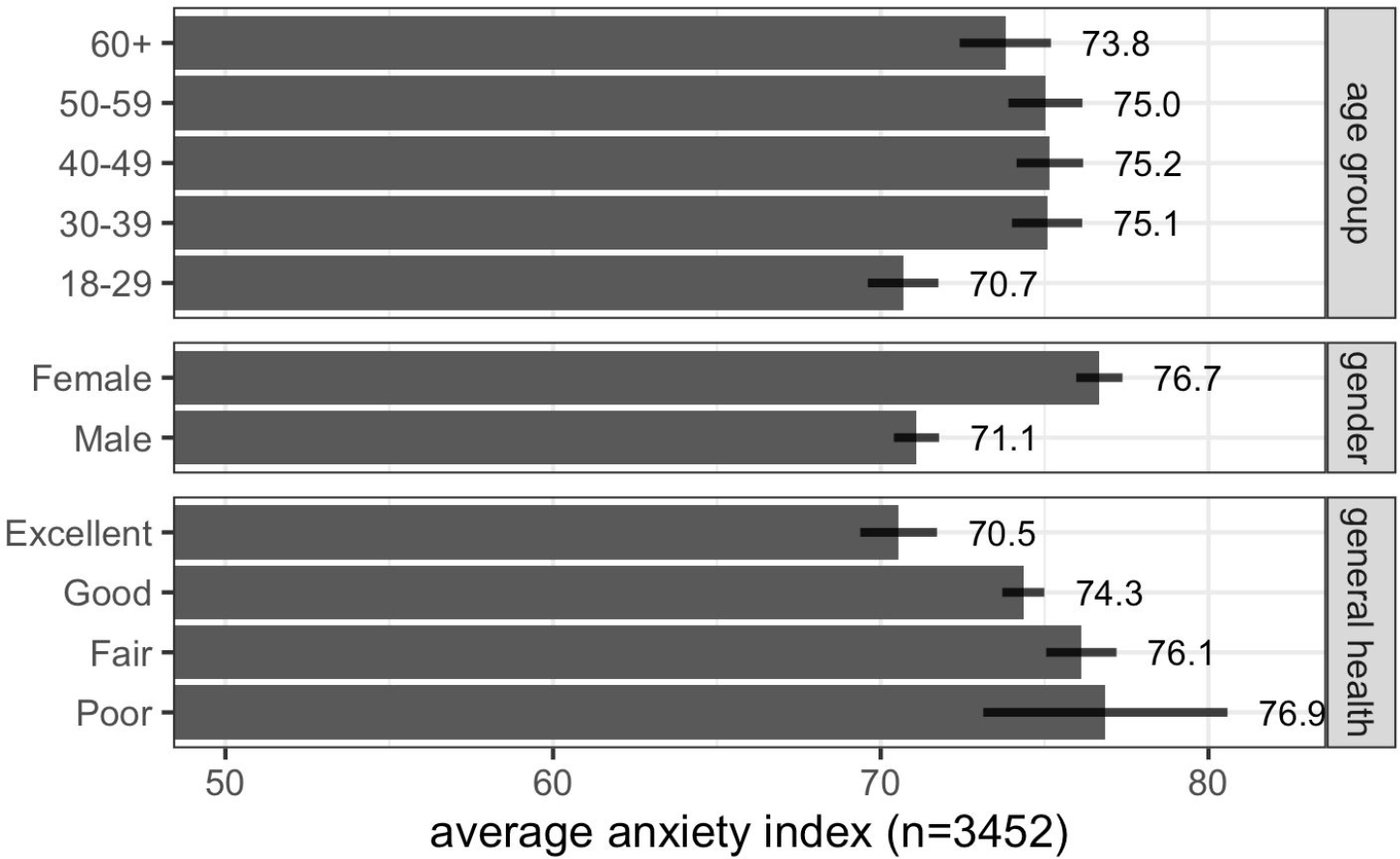
Respondents’ Overall Coronavirus-related Anxiety Level: By Demographic Group. Responses are the average (by demographic group) of 5 responses to 5-point questions: “*I am nervous when I think about current circumstances* (2) “*I am calm and relaxed”* (3) “*I am worried about my health”* (4) “*I am worried about the health of my family members” (5) “I am stressed about leaving my house”*. Averages shown with 95% confidence intervals.

## 6. Nudges to improve attitudes already near their maximum have little effect

In case the messaging was not effective, we implemented a fully randomized survey experiment with informational interventions to try to drive strong pro-health beliefs (Figure 1 and 2) and positive opinions about the crisis response (Figure 5 and 6). A third of respondents assigned to the control group received no message, a third received a message about social externalities with a prompt for reflection (“*If you are infected with COVID-19, you are likely to infect 2,000 people - including your and your friends’ parents and grandparents, who might die. Join the great majority of Italians and stay home. Please write 1-2 sentences reflecting on this message. How does it make you think/feel?”*), and the last third received a message about social externalities with a prompt to name a vulnerable loved one (*“If you are infected with COVID-19, you are likely to infect your parents, your friends’ parents, and/or one any one of your close ones. If they are older than 50, they may die. In the text field below, please name a loved one (example: my father, my grandmother) that you’re most worried about contracting COVID-19*.*”*).

In fact, these beliefs were so close to the ceiling to begin with, there was little room for an intervention to have an effect.

Figure 13 and 14 report the results. Nearly everyone in all three messaging groups – social externalities with reflection, social externalities with personal naming, and no message – agreed on the necessity of canceling social gatherings, closing non-essential businesses, imposing a curfew, and avoiding physical contact. The treatments did little to decrease skepticism or cause backlash (Figure 14). We further did not find that receiving treatments changed any intention to leave home in the coming week or the reasons for doing so. There were also no significant treatment heterogeneities on any outcome by gender, age, health group, anxiety levels, or any past behavioral reports or other attitudinal traits.

This is consistent with a theory of *information overload*: that is, before our intervention, everyone already received this information and accepted its importance from previous studies, advertising, government messaging, friends, family, and other sources (though some rejected). Therefore, additional information is unlikely to change anyone’s beliefs on the necessity of the given measures. Indeed the maximum theoretical effect (which would be seen by moving these percentages up to 100%) was close to the measurement uncertainty.

Finally, were people willing to share this message? We gave respondents the option of sharing a message (of their choosing) with up to 5 people. The vast majority (90%) chose not to share the message, but ∼100 people who received our reflection treatment were significantly less likely to share with anyone. Those who did share were consistently people who strongly believed in the efficacy of social distancing. Nothing else predicted sharing behavior, including receiving an information treatment. There are two explanations for this: (1) most skipped over this prompt (2) most, other than those who strongly endorse social distancing, believe their friends and families already know, and wouldn’t additionally benefit from, this information. The latter is consistent with our theory of *information overload*. It is likely that we can greatly expand this behavior by tuning this message.

Overall, this experiment suggests receiving new information is not changing beliefs, behavioral intentions, or message-spreading, largely because the population had adopted these beliefs prior to our survey. In other words, the control group, with no messaging from us, has been receiving the same messaging in other ways for weeks.

## 7. Conclusions

The Italian population is receiving public health messaging, understands it, and most – save for small numbers of young people and skeptics – claim to be following it closely. Those who take it seriously defect for mostly universally recognized acceptable reasons (e.g., groceries, medicine). New messaging campaigns designed to accomplish the same purpose should not be the focus at this point.

However, the negative psychological effects of the quarantine – most commonly boredom, perceived immobility, and anxiety – are beginning to wear on people and seem likely to become more serious over time. If we expect the Italian people to remain inside and keep following the recommendations of public health officials for the next few weeks or even months (as may be necessary to curb the spread of COVID-19) it would be helpful to find ways of reducing these negative effects of the quarantine. A messaging campaign that suggests how to make the quarantine easier and more frictionless, rather than repeating the now well-known dire reasons to stay indoors, should begin. Possibilities include messaging about online collective exercises, social reading activities (e.g., Perusall.com for schools and universities), safe ways for people to get fresh air outside, classes (e.g., edx.org for online classes), or novel ways of bridging social capital between young and old, or even distribution of inexpensive tablets or laptops to ensure the whole population has video conferencing abilities. If we look for activities and interventions that make the quarantine easier for those affected by it, the population may be able to better tolerate the treatment for as long as it takes to overcome this public health crisis.

## Data Availability

Anonymized replication data is publicly available on a repository at Harvard Dataverse.

https://dataverse.harvard.edu/dataset.xhtml?persistentId=doi:10.7910/DVN/1SBQCX

## Appendix A Statement of Ethical Approval

This work was conducted by the authors in the course of advising the Italian government, which made final decisions about data collection (the government had no prior approval of our paper). Although the government accepted all our advice about the study, we were advised by our IRB that our research formally falls into the category of “not research”.

## Appendix B Survey Information

## Methodology

We asked respondents to report the extent to which they engaged in five officially recommended healthy behaviors in the past week: avoiding social gatherings, informing others of any coronavirus symptoms (if applicable), keeping social distance of 2m from others, staying at home, and washing hands more frequently than usual.

We also ran a survey experiment, randomly assigning potential respondents into three experimental groups. One group received a message about coronavirus communicating possible social externalities of not staying home and asked respondents to reflect on their feelings. The intention behind reflection is focusing the respondents on their emotional response, specifically to the thought of them being responsible for others’ deaths. The message was as follows:

> *“If you are infected with COVID-19, you are likely to infect 2,000 people - including your and your friends’ parents and grandparents, who might die. Join the great majority of Italians and stay home*.

> *Please write 1-2 sentences reflecting on this message. How does it make you think/feel?”*

Another group received a similar social externalities message, but were then asked to name a loved one they believed was particularly susceptible to COVID-19. The intention behind naming is to prime the respondent of an intensely personal impact of non-compliance with the quarantine. The message was as follows:

> *“If you are infected with COVID-19, you are likely to infect your parents, your friends’ parents, and/or one any one of your close ones. If they are older than 50, they may die*.
>
> *In the text field below, please name a loved one (example: my father, my grandmother) that you’re most worried about contracting COVID-19*.*”*

The third group received no message, as a control condition for comparison. In a pre-test, the social externalities messaging outperformed most other messages framing the risks of noncompliance with safety measures - including messages framed around pro-sociality, expert or celebrity advice, and healthcare sector externalities (i.e. “flatten the curve”).

The survey proceeded by then asking respondents for their beliefs, social perceptions, and perceived efficacy about government and/or public safety measures. Respondents were also asked about their trust in government and the degree to which the government was providing factual information about the crisis. We asked respondents a series of questions about anxiety, stress, and worry around the current circumstances.

Next, respondents were asked about their future behaviors - namely whether they planned to leave the house in the next 5 days, and their top reasons for leaving, as well as negative aspects of complying with staying at home.

Finally, respondents took a brief personality battery, provided personal demographic information, and were asked whether they would share either the social externalities message or some other message of their choosing with upto 5 of their friends or loved ones via text or email.

## Survey Details

To draw our sample, we used the market research platform Lucid, which has been shown to produce high quality, representative samples (Coppock and McClellan, 2019). Data was collected between March 18th and March 20th, 2020, with the following characteristics:

- 3452 respondents,
- 1655 males, 1675 females, 14 others, 108 unidentified
- 615 aged 18-29, 732 aged 30-39, 639 aged 50-59, 440 aged 60+

We asked respondents the following questions, most of which came from several members of our team involved in a related effort getting underway (see https://osf.io/3sn2k/).

### 1. Self-Reported Past Behaviors

*In past week, on a scale from 1-100, to what extent did the following describe your behavior?*

- I stayed at home
- I did not attend social gatherings
- I washed my hands more frequently than last month
- I kept a distance of at least two meters to other people
- If I had exhibited symptoms of the disease [COVID-19], I would have immediately informed the people around me

### 2. Information Treatment

We selected the following treatments from a larger set of messages (with a variety of emotional and other appeals) which produced the highest treatment effect on subsequent outcomes on a pre-test of roughly 2000 respondents in the prior week.

**Condition 1: Control**

(no message)

**Condition 2: Externalities message + reflection**

> *If you are infected with COVID-19, you are likely to infect 2,000 people - including your and your friends’ parents and grandparents, who might die. Join the great majority of Italians and stay home*.
>
> *Please write 1-2 sentences reflecting on this message. How does it make you think/feel?*

**Condition 3: Externalities message + naming**

> *If you are infected with COVID-19, you are likely to infect your parents, your friends’ parents, and/or one any one of your close ones. If they are older than 50, they may die*.
>
> *In the text field below, please name a loved one (example: my father, my grandmother) that you’re most worried about contracting COVID-19*.

### 3. Beliefs, Perceptions, Emotions

#### 3.1. Personal beliefs about coronavirus measures

- *Should people in Italy cancel participation in social gatherings because of coronavirus right now?*
- *Should people in Italy not shake other people’s hands because of coronavirus right now?*
- *Should non-essential shops in Italy be closed because of coronavirus right now?*
- *Should there be a general curfew in Italy because of coronavirus right now?*

#### 3.2. Perception of others beliefs about coronavirus measures

- *How many of 100 Italians do you think believe people in Italy should cancel participation in social gatherings because of coronavirus right now?*
- *How many of 100 Italians do you think believe people in Italy should not shake other people’s hands because of coronavirus right now?*
- *How many of 100 Italians do you think believe non-essential shops in Italy should be closed because of coronavirus right now?*
- *How many of 100 Italians do you think believe there should be a general curfew in Italy because of coronavirus right now?*

#### 3.3. Financial sanctioning of risky behaviors

- *How much would you fine participation of social gatherings (out of EUR)?* [0-500]
- *How much would you fine going out despite symptoms of coronavirus (out of EUR)?* [0-500]

#### 3.4. Perceptions of government/public response & efficacy

- *Do you think the reaction of the Italian government to the current coronavirus outbreak is appropriate, too extreme, or not sufficient?* [5 point scale]
- *How much do you trust the Italian government to take care of its citizens?* [5 point scale]
- *How factually truthful do you think the Italian government has been about the coronavirus outbreak?* [5 point scale]
- *Do you think the reaction of the Italian public is appropriate, too extreme, or not sufficient?* [5 point scale]
- *How effective are social distancing measures (e*.*g*., *through a general curfew) to slow down the spread of the coronavirus?* [5 point scale]

#### 3.5. Anxiety/stress/worry battery

- *I am nervous when I think about current circumstances*. [5 point scale]
- *I am calm and relaxed*. [5 point scale]
- *I am worried about my health*. [5 point scale]
- *I am worried about the health of my family members*. [5 point scale]
- *I am stressed about leaving my house*. [5 point scale]

### 4. Future Behaviors

*Do you need to leave home in the next 5 days?* [y/n]

*What are the reasons personally for you to leave your home (check all that apply)?*

- Going to work
- Walking a pet
- Doing physical activity (e.g. exercising, jogging)
- Procuring food for yourself or family
- Going to the pharmacy
- Going to the hospital / receiving medical treatments
- Taking care of dependents
- Meeting friends or relatives
- Getting tired of being inside of the house
- Getting bored
- Getting some adrenaline (from breaking the law)
- Exercising my freedom
- Other

*What are the main negative sides of complying with stay home requirement (check all that apply)?*

- Conflicts in the family
- Boredom
- Lack of exercise
- Lack of fresh air
- Lack of freedom
- Loss of job / income source
- Homeschooling
- Lack of social activities
- Combining homeschooling and working on distance
- Loneliness
- Other

### 5. Personality Battery

- *To which extent do the following questions apply to you? I see myself as …*
- Extraverted, enthusiastic [5 point scale]
- Critical, quarrelsome [5 point scale]
- Dependable, self-disciplined [5 point scale]
- Anxious, easily upset [5 point scale]
- Open to new experiences, complex [5 point scale]
- Reserved, quiet [5 point scale]
- Sympathetic, warm [5 point scale]
- Disorganized, careless [5 point scale]
- Calm, emotionally stable [5 point scale]
- Conventional, uncreative [5 point scale]

### 6. Personal Info

- *What year were you born?* [1919-2020]
- *Which gender do you identify with?* [Male/Female/Other]
- *What is your postal code?*
- *How healthy are you?* [poor/fair/good/excellent]
- *How many of the following conditions do you have: cardiovascular diseases, diabetes, hepatitis B, chronic obstructive pulmonary disease, chronic kidney diseases, and cancer?* [1-5]

### 7. Sharing

*In order to help Italy in the fight against the current Coronavirus epidemic, will you share one of the following messages below with 5 of your colleagues, friends, or loved ones? If so, select one (or send the default one selected) and please enter their email addresses or mobile numbers below*.

- If you are infected with Coronavirus, you are likely to infect 2,000 people - including your and your friends’ parents and grandparents, who might die. Join the great majority of Italians and stay home. [**externalities**]
- If you are infected with Coronavirus, you are likely to infect your parents, your friends’ parents, and/or one any one of your close ones. If they are older than 50, they may die. In the text field below, please name a loved one (example: my father, my grandmother) that you’re most worried about contracting Coronavirus. [**externalities + reflection**]
- The vast majority of Italians believe that all Italians should stay home. Join them in the fight against the Coronavirus and stay home. [**majority**]
- Staying at home does not need to be boring! Call friends regularly, exercise, and try out new hobbies --- these are some of the things that will make your time at home more pleasant. Stay home. [**fun**]
- The doctors and nurses who are risking their lives to fight the Coronavirus are urging all Italians to stay home. The health system is struggling. Stay home. [**experts**]

## Footnotes

3 Translated from original Italian via Google Translate: *“Credo che TUTTI dovrebbero rimanere a casa, uscendo solo per lo stretto necessario e adottando le misure di sicurezza. Solo cosí si può arginare e risolvere questo enorme problema nel minor tempo possibile*.*”* Representative reflection was assessed from a random sample of reflection texts.

4 Translated from original Italian via Google Translate: *“Capisco perfettamente tutto quello che sta accadendo, quindi mio malgrado, resto in casa, ma vorrei che questo lo comprenda tutta quella gente che purtroppo continua a uscire, soprattutto i giovani, anche tutti i giorni! Penso che più resteremo a casa, prima ne usciremo!”*

5 We define skeptics as those reporting opinions reflecting skepticism or distrust of government, government information, public health protocols, or the severity of the COVID-19 crisis. This definition classifies those who say the response is “extreme” as skeptics, but not those who say the response is “insufficient”.

6 Translated from original Italian via Google Translate: *“Io non credo. È tutto molto mediatico. Anche l’anno scorso 12 mila morti di influenza e nessuno ne ha parlato. Comunque eseguo gli ordini anche se non li capisco. Sicuramente non sta comandano Conte ma qualcun altro*.*”*

